# Study of pre-hospital care of Out of Hospital Cardiac Arrest victims and their outcome in a tertiary care hospital in India

**DOI:** 10.1101/2020.06.12.20129502

**Authors:** Rachana Bhat, Prithvishree Ravindra, Ankit Kumar Sahu, Roshan Mathew, William Wilson

## Abstract

**BACKGROUND:** India does not have a formal cardiac arrest registry and a centralized emergency medical system. In this study, we aimed to assess the prehospital care received by the patients with OHCA and to predict the factors that could influence their outcome.

**METHODS:** We performed a prospective observational study, including OHCA patients presenting to the emergency department (ED) between February 2019 and January 2020. A structured proforma was used to capture information like basic demography, prehospital details like bystander cardiopulmonary resuscitation (CPR), use of an automated external defibrillator (AED), clinical profile, and outcome.

**RESULTS:** Among the included 205 patients, the majority were male (71.2%) and belonged to older age (49.3%). The nature of arrest was predominantly non-traumatic (82.4%). The initial rhythm at presentation was non-shockable (96.5%). Return of spontaneous circulation (ROSC) was achieved in 17 (8.3%) patients, of which only 3 (1.4%) patients survived till discharge. The home was the most common location of OHCA (116, 56.6%). Among the OHCA patients, witnessed arrests were seen only in 64 (31.2%), of which 15 (7.8%) received bystander CPR, and AED was used in 1% of the patients. The initial shockable rhythm was a significant predictor of ROSC (OR 18.97 95%CI 3.83-93.89; p<0.001) and survival to discharge (OR 42.67; 95%CI 7.69-234.32; p<0.001).

**CONCLUSION:** In a developing country like India, this study underlines the poor status of the prehospital system like lower bystander CPR, AED and ambulance usage. Moreover, ROSC was seen only in less than 10% of patients, and only 1.3% got discharged from the hospital.

## Introduction

The global incidence of out of hospital cardiac arrest (OHCA) is estimated to be 55 per 100,000 person-years^1^, making it an important public health challenge. The data from India regarding OHCA is lacking as there is no national cardiac arrest registry at present. The challenges pertaining to OHCA in India are unique and multifaceted. The awareness among the community regarding cardiopulmonary resuscitation (CPR) and AED is low. ^2,3^ Bystander CPR rate has been found to be 1.3%^4^, much lower than most countries. The emergency medical services (EMS) are fragmented and not accessible throughout the country. Most ambulances are used as a transport vehicle to reach the hospital, without simultaneously delivering emergency care in the ambulance. The Emergency Medicine paramedic courses have been initiated in a few institutes in the last decade, though a robust system of EMS is yet to be developed.

Our study objective was to assess the prehospital care received by the patients presenting to the Emergency Department with OHCA in a tertiary care hospital in India and to predict the factors that could influence their outcome.

## Methods

The study was a prospective observational study conducted at a tertiary care centre in India. This study was started only after waiver of ethical approval from Institutional Review Board, Kasturba Medical College, Manipal, Manipal Academy of Higher Education, Manipal, Karnataka. Every patient presenting to the hospital with OHCA during the period from Feb 2019 to January 2020 were considered for the study. Cases with age less than 18 years or where the legally acceptable representative denied consent for the study were excluded. The primary objective of the study was to assess the pre-hospital care received by victims of OHCA including bystander CPR, EMS utilisation & interventions, AED use. The secondary objective was to assess the factors influencing ROSC and survival to hospital discharge.

After informed consent from the legally acceptable representative, details of patients presenting to the Emergency Department with OHCA were collected. The patients attaining ROSC were followed up to discharge/death. CPC was calculated at discharge to assess the neurological outcome. During the course of hospital stay, details such as initiation of targeted temperature management, time to revascularization (if applicable), final diagnosis was noted.

The data was entered in MS EXCEL spreadsheet and analysis was done using SPSS (version 23; IBM, Armonk, NY). Categorical variables were presented in number and percentage (%) and continuous variables were presented as mean (standard deviation) and median (interquartile range). Logistic regression analyses were used to predict the effect of different variables on the ROSC, 24-h survival, and survival to discharge. The variables included in the logistic regression model were age, gender, arrival time to hospital, witnessed arrest, By-stander CPR received, duration of cardiac arrest, traumatic/non-traumatic cardiac arrest and type of rhythm (shockable or non-shockable). A p value less than 0.05 was considered statistically significant.

## Results

The study included 205 patients with OHCA presenting to the Emergency Medicine department of a tertiary care hospital. The Modified Utstein template is provided in Figure 1. 56.6% (n=116) of the patients sustained cardiac arrest at home and 30.7% (n=63) of the patients sustained cardiac arrest in transit. The demographic characteristics, comorbidity profile and the initial presenting symptoms of the patients are provided in Table 1.

**Table 1:** Basic characteristics of the included patients.

|  |  |
| --- | --- |
| Characteristics |  |
| **Age<sup>1</sup>** | 58.4(17.1) |
| **Age Group<sup>2</sup>** |  |
| 18-39 years | 32 (15.6%) |
| 40-59 years | 72 (35.1%) |
| > 59 years | 101 (49.3%) |
| **Gender<sup>2</sup>** |  |
| Male | 146 (71.2%) |
| Female | 59 (28.8%) |
| **Initial presentation<sup>2</sup>** |  |
| Unresponsive | 64 (31.2%) |
| Breathlessness | 37 (18%) |
| Trauma | 36 (17.6%) |
| Chest pain | 24 (11.7%) |
| Giddiness/ syncope | 18 (8.8%) |
| Fever | 6 (2.9%) |
| Bleeding | 4 (2%) |
| Hanging | 4 (2%) |
| Headache | 3 (1.5%) |
| Abdominal Pain | 3 (1.5%) |
| Drowning | 3 (1.5%) |
| Poisoning | 2 (1%) |
| Electrocution | 1 (0.5%) |
| **Co-morbid illness<sup>2</sup>** |  |
| Hypertension | 71 (34.6%) |
| Diabetes | 59 (28.8%) |
| Coronary artery disease | 38 (18.5%) |
| Asthma/COPD | 14 (6.8%) |
| Cerebrovascular accident | 11 (5.3%) |
| Chronic Kidney disease | 7 (3.4%) |
| Heart failure | 7 (3.4%) |
| Malignancy | 5 (2.4%) |
| Others<sup>3</sup> | 9 (4.3%) |
| **Type of arrest<sup>2</sup>** |  |
| Non-traumatic cardiac arrest | 169 (82.4%) |
| Traumatic cardiac arrest | 36 (17.6%) |

1. Mean (SD)
2. N (%)
3. **Others:** Chronic liver disease (2), Seizure (1), Deep vein thrombosis/Pulmonary embolism (1), Hypertrophic cardiomyopathy (1)

**Figure 1:**
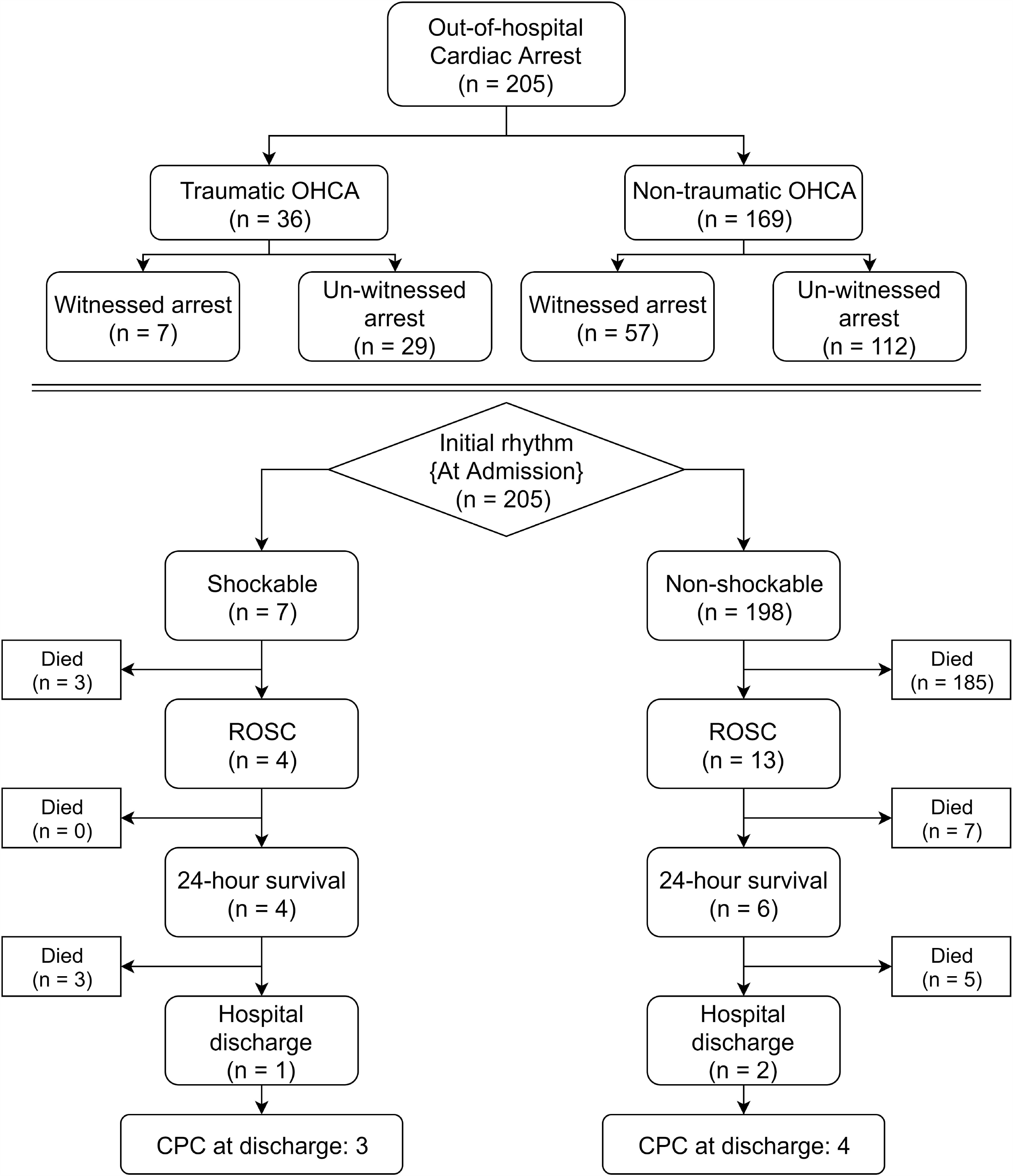
Modified Utstein template OHCA: Out of hospital cardiac arrest; ROSC: Return of Spontaneous circulation; CPC: Cerebral performance category

Pre-hospital factors such as place of arrest, mode of transport to hospital and bystander CPR were analysed and details mentioned in Table 2. A significant percentage of our patients (41.5%) reached the hospital by means other than the ambulance. Of the available data, median ambulance response time was 10(IQR: 10-20) minutes. It is worthy to note that only 6.8% (n=14) patients received bystander CPR. Among the patients transported in the ambulance only 7.8% (n=15) had received CPR. AED was used only in one percent of the patients. No patients had received prehospital shocks during resuscitation or achieved prehospital ROSC.

**Table 2:** Pre-hospital response system.

|  |  |
| --- | --- |
| **Place of arrest** |  |
| Home | 116 (56.6%) |
| In transit | 63 (30.7%) |
| Road | 22 (10.7%) |
| Workplace | 2 (1%) |
| Railway station | 1 (0.5%) |
| Restaurant | 1 (0.5%) |
| **Pre hospital response** |  |
| Witnessed | 64 (31.2%) |
| By-stander CPR | 20 (9.8%) |
| AED used | 2 (1%) |
| **Mode of arrival to study centre** |  |
| Ambulance | 120 (58.5%) |
| Personal vehicle | 76 (37.1%) |
| Three-wheeler | 8 (3.9%) |
| Bus | 1 (0.5%) |
| **Who Brought** |  |
| **Bystander** | **185 (90.2%)** |
| Relatives | 162(79%) |
| Colleagues/Friends | 14(6.8%) |
| Civilian | 9(4.4%) |
| **First responders** | **20 (9.8%)** |
| Paramedics | 14(6.8%) |
| Police | 4 (2%) |
| Fire Rescuer | 2 (1%) |

Among the patients with witnessed arrest (31.2%, n=64), the median time of cardiac arrest before arrival to our hospital was 30 (IQR: 15 -41.25) minutes. With respect to the initial rhythm on presentation, shockable rhythm was seen in seven patients (3.4%); ventricular fibrillation in 6 patients and pulseless ventricular tachycardia in one patient. 96.6% (n=198) had non shockable rhythm of which 13 patients (6.3%) had pulseless electrical activity (PEA). Return of Spontaneous Circulation (ROSC) was achieved in 17 (8.3%) patients. The final diagnoses of patients based on clinical gestalt and post-mortem reports are provided in Figure 2. Amongst the patients who achieved ROSC, ten patients(4.8%, 10/205) survived more than 24 hours and only three(1.4%) could survive till hospital discharge of which one had a cerebral performance score of 3 and two had a score of 4. Four out of seven patients (57%; p=0.001) with initial shockable rhythm achieved ROSC as compared to 13 out of 185 (7%) with non-shockable rhythm (p=0.001).

**Figure 2:**
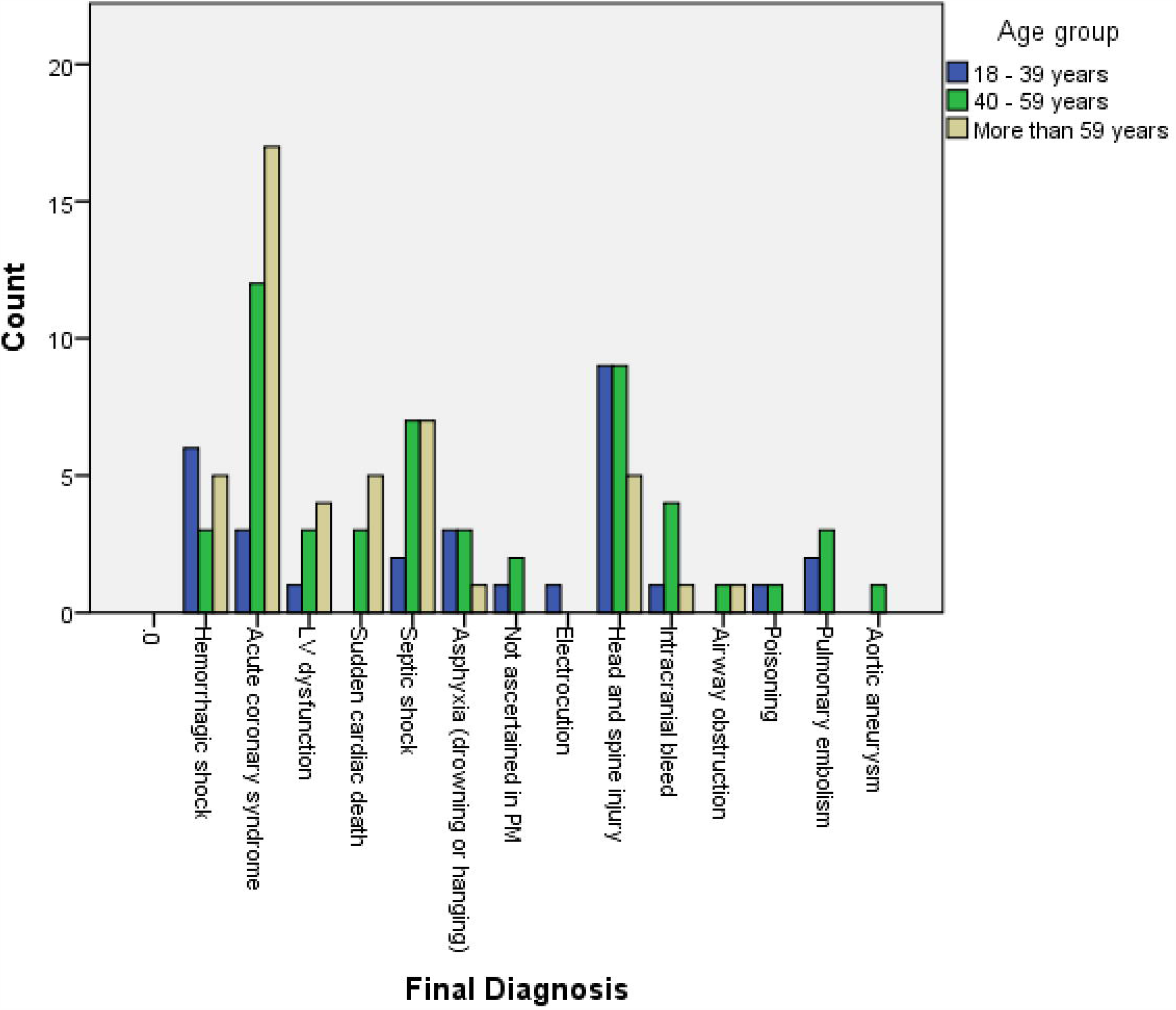
Final diagnoses of patients of OHCA

Factors associated with ROSC, survival to hospital discharge and favourable neurological outcome at discharge were analysed with logistic regression analysis and it was found that the duration of cardiac arrest and shockable rhythm at presentation had a statistically significant association with ROSC (Table 3). Non-traumatic cardiac arrest had a higher incidence of ROSC than traumatic cardiac arrest, although it was not statistically significant. Longer the duration of cardiac arrest, lower the odds of ROSC (OR 0.937; 95% CI 0.88-0.99; p-0.047) whereas initial shockable rhythm was associated with higher odds of survival (OR 18.97 95%CI 3.83-93.89; p<0.001). Similarly, patients with initial shockable rhythm had statistically significant and higher odds of survival for more than 24 hours of ROSC (OR 42.67; 95%CI 7.69-234.32; p<0.001).

**Table 3:**
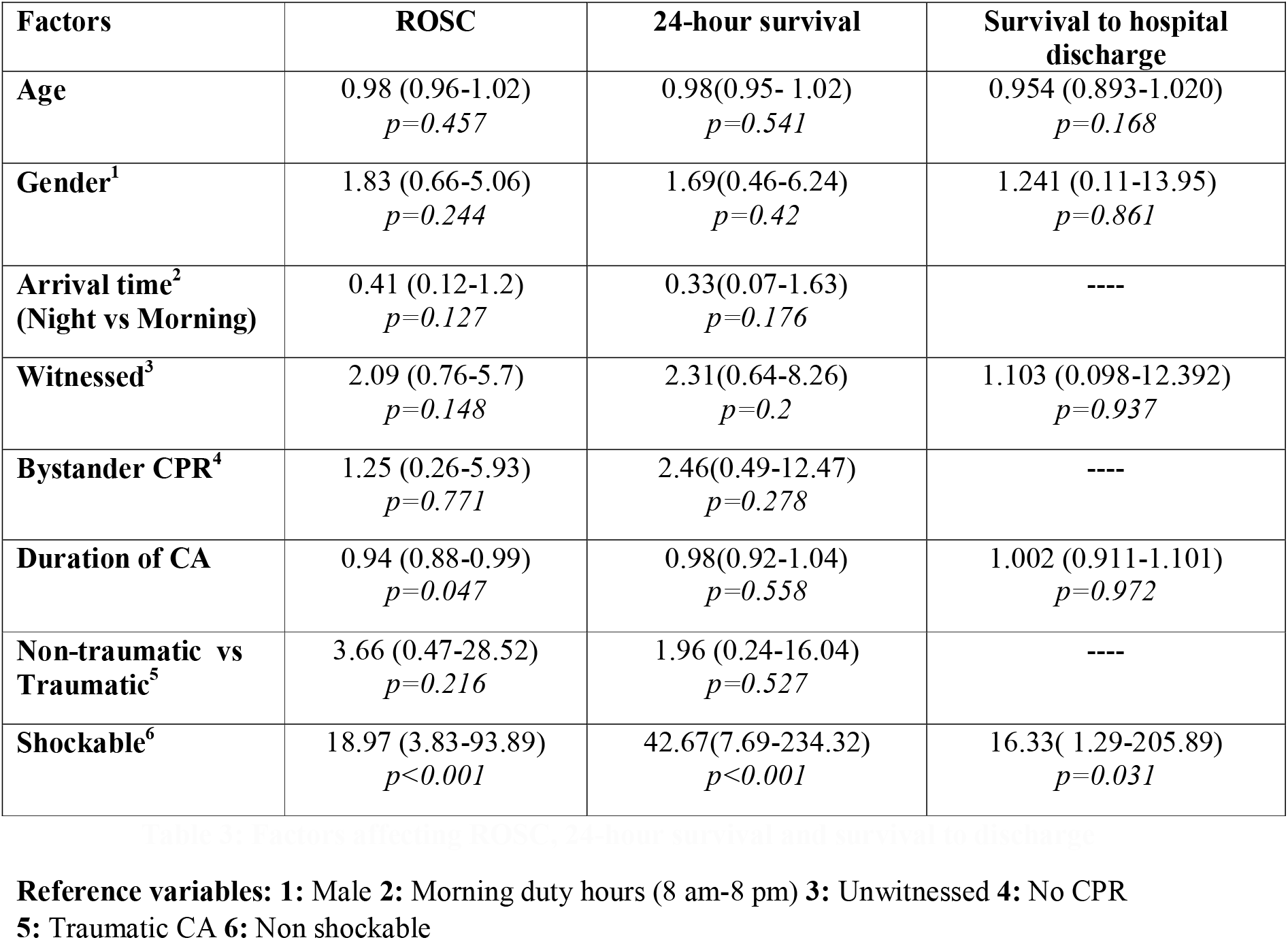
Factors affecting ROSC, 24-hour survival and survival to discharge.

## Discussion

Our study aimed to understand the pre-hospital care received by the OHCA victims and factors that could influence ROSC in a developing country like India. We report that the majority of OHCA happened at home (56.6%); however a significant proportion (30.8%) happened during transit (24.9% in ambulances, 5.9% in private vehicles). This is in line with the findings of the Cardiac Arrest Resuscitation Outcome (CARO) study which had 92% of the arrests at home.^4^ The Swedish Cardiac arrest registry also found that the majority of older adults suffered a cardiac arrest at home followed by in transit (ambulance) to the hospital.^5^ In our study we noted that a significant percentage (41.4%) of cardiac arrest victims were transported in means other than ambulance. Wijeshkara et al found that only 1.4% to 19.4% of ED patients utilised EMS to get to the ED.^6^ A systematic review & meta-analysis by Yan et al showed that prehospital stage was the most impactful stage for care of OHCA patients and where efficient CPR from EMS translated to highest incidence of ROSC (36.3%; 95% CI 23.8 - 48.9%).^7^ This emphasizes the need to educate the community regarding the importance of using an ambulance and EMS systems, so that earlier AED use and ACLS interventions can be administered.^8^

Continuing with the quality of the EMS services available to our patient group only 7.8% received CPR, with only 12.5% of ambulances having Basic Life Support (BLS) trained personnel. Ramanujam et al in a single centre study in Chennai found that 85% of the trauma victims did not receive any formal prehospital interventions, further emphasizing the lack of prehospital EMS facilities available although in a different patient group.^9^ Our study also found that the AED use was limited and there were no prehospital shocks administered or prehospital ROSC. This underlines the fact that ambulances are being used as mere transport vehicle rather than a device to deliver pre-hospital care. BLS ambulance of >4 per neighbourhood and Advanced Life Support (ALS) ambulances of >1.5 per neighbourhood have been linked with successful ROSC.^10^ Considering that a significant proportion of cardiac arrests are happening in the ambulance during transit, it is essential to realise the importance of having trained EMS personnel in fully BLS and ALS equipped ambulance and develop a stronger EMS system in the country.

It is well established that by-stander CPR is one of the cornerstones in improving OHCA results with better survival and discharge rates as compared to no bystander CPR.^11,12^ Amongst the sparse data available from India, the CARO study reported bystander CPR rate of 1.3%^4^ Our study found that bystander CPR was done only in 6.8% of our patients which falls way below the impact goal of bystander response of 62% set by the AHA-ECC.^13^The recent Sweden registry which reported bystander CPR rates of 26-54% across the adult age groups.^5^ To bridge this gap, we need to increase the knowledge in the community about CPR by having widespread training programs throughout the country aimed at dispelling myths and educational campaigns and also initiate school based training.^14^

Among the available final diagnoses assessed of the OHCA victims, majority had cardiovascular cause. This is consistent with studies elsewhere showing cardiovascular cause as the leading diagnoses among the victims.^15^ In our study, 15.6% had clinical/ post-mortem final diagnosis as acute coronary syndrome. It has been shown that reduced transport times may benefit patients with OHCA and STEMI also translating to significantly better neurological outcomes.^16^ This emphasises the need for transport using the EMS with an AED with BLS/ALS trained personnel, along with STEMI alert system. Haemorrhagic shock was the cause of death for 6.8% of the patients, which also included isolated limb injuries. The resuscitation of these patients in transit including simple measures such as compression of the bleeding site and fluid resuscitation may have improved outcome.

We found that 8.3% had ROSC, 4.9% had 24-hour survival and 1.4% survived to discharge. The factors positively influencing outcome included initial shockable rhythm and shorter duration of arrest. None of the discharged patients had favourable CPC (CPC 1, 2). CARO study found that 32.5% had ROSC, 8.8% had survival to discharge and 3.8% had favourable CPC. They concluded that there is a need for creating a centralised medical emergency body to oversee the setting up of EMS.^4^ This re-emphasises the importance of community training to initiate early CPR, stronger EMS system, early AED use to improve the outcomes of OHCA in India.

## Conclusion

The pre-hospital care received by the OHCA victim in India needs immediate attention. Low by-stander CPR rate, under-utilised and under-equipped EMS system are the challenges that we face. Community education and training programs along with strengthening the EMS systems are the way forward to improve care of OHCA victims in India.

### Limitations

As the number of patients who had 24 hour survival and survived to discharge were less, the study may not have been able to detect all the factors influencing these outcomes. The information such as ambulance response time was missing in few cases.

## Data Availability

available in manuscript

## CREDIT authorship contribution statement

Conceptualisation: RB, PR

Data curation: WW

Formal analysis: AS, RM

Investigation: RB, PR, WW

Methodology: RB

Software: AS, RM

Supervision: RB, PR

Visualisation: AS

Writing – original draft: RM, WW, PR

Writing – review and editing: RM, RB

